# Prevalence and determinants of cervicovaginal, oral, and anal HPV infection in a population of transgender and gender diverse people assigned female at birth

**DOI:** 10.1101/2023.08.15.23294129

**Authors:** Ryan D. McIntosh, Emily C. Andrus, Heather M. Walline, Claire B. Sandler, Christine M. Goudsmit, Molly B. Moravek, Daphna Stroumsa, Shanna K. Kattari, Andrew F. Brouwer

**Affiliations:** Department of Epidemiology, University of Michigan, Ann Arbor, MI, United States; Department of Otolaryngology, University of Michigan, Ann Arbor, MI, United States; Reproductive Endocrinology Clinic, Center for Reproductive Medicine, University of Michigan, Ann Arbor, MI, United States; School of Social Work, University of Michigan, Ann Arbor, MI, United States; Department of Women’s and Gender Studies, University of Michigan, Ann Arbor, MI, United States

**Keywords:** HPV, transgender, gender diverse, cervicovaginal, anal, oral, self-sampling

## Abstract

**Introduction:** HPV causes oral, cervicovaginal, and anogenital cancer, and cervical cancer screening options include HPV testing of a physician-collected sample. Transgender and gender diverse (TGD) people assigned female at birth (AFAB) face discrimination and stigma in many healthcare settings; are believed to be a lower risk for cervical cancer by many physicians; are less likely to be up to date on preventive health care services such as pelvic health exams; and are more likely to have inadequate results from screening tests. Self-sampling options may increase access and participation in HPV testing and cancer screening.

**Methods:** We recruited 137 TGD individuals AFAB for an observational study, mailing them a kit to self-collect cervicovaginal, oral, and anal samples at home. We tested samples for HPV genotypes 6, 11, 16, 18, 31, 33, 35, 39, 45, 51, 52, 56, 58, 59, 66, 68, 73 and 90 using a PCR mass array test.

**Results:** 102 participants completed the study. Among those with valid tests, 8.8% were positive for oral HPV, 30.5% were positive for cervicovaginal HPV, and 39.6% were positive for anal HPV. A large fraction of anal (50.0%) and oral (71.4%) infections were concordant with a cervicovaginal infection of the same type.

**Conclusions:** HPV infection in TGD people AFAB may be just as high, if not higher, than in cisgender women. It is essential that we reduce barriers to cancer screening for TGD populations, such as through the development of a clinically approved self-screening HPV test.

## Introduction

The human papillomavirus (HPV) causes more than 24,000 cancers each year in women and people assigned female at birth (AFAB), including 12,000 cervical cancers [1]. The U.S. Preventive Services Task Force (USPSTF) recently revised cervical cancer screening guidelines to include the option of testing for high-risk HPV instead of, or in addition to, cytological (Pap) screening [2]. Many physicians believe transmasculine people (those who were AFAB but who identify as male or masculine [3]) and nonbinary and gender diverse people (those who identify between or beyond the male–female gender binary; note that transgender and gender diverse may be overlapping identities) to be at a lower risk for cervical cancer than cisgender (i.e., non-transgender) women because of lower rates of penile–vaginal intercourse [4]. However, prior studies have suggested that transmasculine populations have just as high or higher rates of sexually transmitted infections and may be at equal or higher risk of HPV-associated cancers [3, 5]. Moreover, transgender and gender diverse (TGD) people face discrimination and stigma in health care settings and daily life and consequently are less likely to seek reproductive and preventive health care services, including cervical cancer screening [6–12]. Recent studies have proposed HPV self-sampling as an alternative to provider-based testing, circumventing some of these known barriers to screening [13–16]. However, any clinically approved self-sampling protocol should be designed to address the specific physical and mental discomfort that TGD people may face. Ultimately, more work is needed to understand the risk of HPV-associated cancers in TGD populations [17] as well as the impact and feasibility of HPV self-sampling as a cervical cancer screening tool. This work is especially crucial in the context of the updated USPSTF guidelines and the current politicization of TGD medical care.

While HPV is best known as the etiological agent of nearly every cervical cancer, it also causes over 90% of anal cancers, 40% of vaginal, vulvar, and penile cancers, and more than 60% of oropharyngeal and other oral cancers [1, 18, 19]. Oropharyngeal cancer is now the HPV-associated cancer with the highest incidence in the U.S., with approximately 19,000 of the 43,000 HPV-associated cancers diagnosed occurring at oropharyngeal sites (approximately 12,000 are cervical, 7,000 are anal, and the rest are at other genital sites) [20]. There is increasing recognition that HPV must be considered a multisite (cervicovaginal, oral, anal) infection to fully understand its epidemiology [21–26]. We extend this multisite perspective to HPV-associated cancer risk, screening, and prevention for TGD people AFAB. Specifically, we enrolled a sample of TGD AFAB people and mailed them at-home, self-sampling kits, which we tested for cervicovaginal, oral, and anal HPV.

## Methods

### Ethics statement

This study was approved by the University of Michigan Institutional Review Board (IRBMED: HUM00166980) and Rogel Cancer Center Protocol Review Committee. Prior to beginning the study, ten TGD community consultants reviewed and commented on the study protocol, surveys, and sample collection instructions; each person was compensated with a $100 gift card for their time. In the main study, all participants gave written, informed consent virtually, using the SignNow platform. Participants were compensated with a $50 gift card for full study completion and were returned the results of their high-risk HPV test, with appropriate cautions given the non-clinical sampling. This study included the use of a cervicovaginal swab outside of the Food and Drug Administration (FDA) approved context (i.e., used by the participant, not by a physician, to collect a sampling for HPV testing); the FDA determined the study, including return of qualified research results to participants, to be a non-significant risk device study.

## Study population

### Eligibility criteria

To participate in the study, potential candidates had to meet all of the following criteria: (1) be between the ages of 21–65, (2) have been assigned female at birth (AFAB), (3) have a male, masculine, nonbinary, queer, or another gender diverse identity other than female or feminine, (4) not be pregnant, (5) not have had cervix removed, (6) not be menstruating on the day of the cervicovaginal swab self-collection, and (7) live in Michigan, Illinois, Indiana, Ohio, or Wisconsin. Individuals of any HPV vaccination status or prior HPV infection status were eligible.

### Recruitment and enrollment

Recruitment methods included advertisements and listserv emails through Michigan Medicine’s Comprehensive Gender Services Program, the University of Michigan Health Research posting site, social media posts, outreach to statewide LGBTQ+ groups and organizations, and word of mouth referrals. Interested participants reached out to study staff, who screened them for eligibility requirements. Potential participants were then invited to an individual virtual video call to review the consent form and details of the study.

### Surveys

Participants completed two surveys on an online platform (Qualtrics). The demographic and behavioral survey include questions on sociodemographic characteristics; healthcare utilization; sexual behavior and history; and substance use. The self-sampling survey, which was administered after specimen collection, included questions on the acceptability, comfort, and ease-of-use of the cervicovaginal and anal self-sampling protocols, as well as preference for self- or physician-administered testing/screening.

### Biospecimen collection and processing

Participants were mailed a study test kit that included materials and instructions for self-collection of 3 separate biospecimens: a cervicovaginal swab, an oral rinse, and an anal swab. Self-sampling instructions were modified to remove gendered language (e.g., “genital swab” or “front hole swab” instead of “cervicovaginal swab” or “Evalyn Brush”). All samples were repackaged in USPS biospecimen compliant packaging then returned via mail to the study. The estimated time lapse between sample collection and package receival was 5 days.

#### Cervicovaginal swab

Participants used an Evalyn Brush (Rovers Medical Devices BV, Oss, Netherlands) to self-collect a cervicovaginal sample [27]. Participants were provided with self-collection instructions that were modified from the original, with permission from Rovers Medical Devices, to remove gendered language. In short, participants were instructed to use the plunger on the device to inserted the brush into their vagina to the appropriate depth, rotate the brush 5 times, and then retract and cap the device for transport. HPV DNA on a dry-stored Evalyn Brush has previously been shown to be analytically stable [28]. Upon return to the study, the protective cap was removed, and the brush tip was vigorously agitated then gently mashed in a 20mL vial of PreservCyt solution (Hologic, Inc.; Marlborough, MA). Since the genital mucus sample often dried during transport, the brush bristles arrived firmly adhered together. Mashing ensured complete separation of the plastic brush bristles, which improved elution of the cells from the brush into the PreservCyt media. The tip of the brush was then removed from the barrel of the device using a set of autoclaved disposable forceps provided by the manufacturer and left to soak for a minimum of 5 minutes to rehydrate dried mucus that would have otherwise impeded cell elution. The device barrel was discarded, but the brush tip was retained within each PreservCyt vial. After resting, each vial was vortexed for 30 seconds to remove cervical cells that were adhered to the brush bristles. A 10mL aliquot was pipetted into a conical tube and sent to our research laboratory for genotyping, a 2mL aliquot was pipetted into a 5mL transport tube and sent to Michigan Medicine Pathology for Cobas testing. Samples were stored at 0^◦^C.

#### Anal swab

Participants were sent a Dacron flocked swab (Puritan Medical Products; Guilford, ME), a 10ml conical tube with 1 mL of DNA/RNA Shield (Zymo Research; Irvine, CA) or Preserv-Cyt. Sample collection instructions were based on those developed by Meites et al. [29]; briefly, participants were instructed to insert the swab 2 inches into anus, twirl the swab between the thumb and forefinger while pressing laterally against the anal walls, and then remove the swab from the body. The swab was then placed in the corresponding conical tube. Upon return to the study, the conical tube containing the Dacron flocked swab and transport media were vortexed for 15 seconds to release cells from the fibers of the swab tip. The swab was then extracted from the tube with a pair of sanitized hemostats and discarded. The sample was sent to our research laboratory for genotyping.

#### Oral rinse

Participants received 1 50 mL conical tube with 10 mL Scope Mouthwash (Proctor & Gamble; Cincinnati, OH). Participants were instructed to swish the mouth wash vigorously for 30 seconds, then spit the contents back into the original 50 mL tube. Upon return to the study, samples were transferred to our research laboratory for genotyping.

### HPV testing

#### High-risk HPV testing

Cervicovaginal samples were tested by Michigan Medicine Pathology clinical laboratories using a Cobas high-risk HPV test (Roche Molecular Systems, Inc; Branchburg, NJ), a polymerase chain reaction (PCR)-based test that provides pooled results on high-risk genotypes and individual results for HPV-16 and HPV-18, the highest-risk genotypes. The results of this test were returned to participants.

#### PCR Mass Array HPV genotyping

Cervicovaginal, anal, and oral samples were tested for HPV genotypes 6, 11, 16, 18, 31, 33, 35, 39, 45, 51, 52, 56, 58, 59, 66, 68, 73 and 90. Sample processing and DNA extraction details are the same as in Eisenberg et al. [30], and technical details of the PCR Mass Array test are given in Walline et al. [31]. Participants whose samples contained insufficient DNA or otherwise resulted in inconclusive test results were denoted as invalid. The results of the PCR Mass Array were not returned to participants.

## Results

### Demographics, behaviors, and medical history

We enrolled 137 participants between August 2020 and August 2021, and 102 participants completed the biospecimen collection and demographic and behavior questionnaire. The demographic, behavioral, and medical history characteristics of these participants are summarized in Table 1; a table summarizing participants’ numbers of recent sexual partners is given in the supporting information (Table S1). While participants ranged from 21 to 59 years of age, the majority of those who completed the study (66%) were under 30 years old. The study participants were predominantly White (82%). Participants were also highly educated, with 92% reporting attending at least some college and 65% having a 2-year, 4-year, or professional/doctorate degree. Reported annual income was distributed fairly evenly between the categories ranging from less than $10,000 to at least $50,000. Most participants reported having a single partner, either married/partnered (46%) or not (12%); about 25% of participants were never married or partnered.

**Table 1:** Demographic, behavioral, and medical history characteristics of participants. N is the number of participants answering the question, and n is the number of respondents answering in the affirmative.

|  | n | % |
| --- | --- | --- |
| <u>Age Group (N=102)</u> |  |  |
| 21-29 | 67 | 65.7% |
| 30-39 | 29 | 28.4% |
| 40-59 | 6 | 5.9% |
| <u>Race/Ethnicity (N=102)</u> |  |  |
| White | 84 | 82.4% |
| Multiracial | 8 | 7.8% |
| Asian | 5 | 4.9% |
| Black | 5 | 4.9% |
| <u>Gender (N=102)</u> |  |  |
| Nonbinary, genderfluid, agender | 49 | 48.0% |
| Male, transgender male, transmasculine | 50 | 49.0% |
| Other / Prefer not to label | 3 | 2.9% |
| <u>Sexual Orientation (N=102)</u> |  |  |
| Bisexual, pansexual, or omnisexual | 44 | 43.1% |
| Queer | 37 | 36.3% |
| Homosexual | 9 | 8.8% |
| Asexual or demisexual | 8 | 7.8% |
| Straight or heterosexual | 3 | 2.9% |
| Other / Prefer not to label | 1 | 1.0% |
| <u>Highest Level of Education (N=102)</u> |  |  |
| High school or less | 8 | 7.8% |
| Some college | 27 | 26.5% |
| 2- or 4- year degree | 46 | 45.1% |
| Professional or doctorate degree | 21 | 20.6% |
| <u>Current Employment Status (N=102)</u> |  |  |
| Full time | 38 | 37.3% |
| Student | 22 | 21.6% |
| Part time | 15 | 14.7% |
| Unemployed, looking for work | 12 | 11.8% |
| Disabled and not working | 10 | 9.8% |
| Unemployed, not looking for work | 5 | 4.9% |
| <u>Yearly Household Income (N=102)</u> |  |  |
| Less than \$10,000 | 17 | 16.8% |
| \$10,000 - \$19,999 | 19 | 18.8% |
| \$20,000 - \$39,999 | 20 | 19.8% |
| \$40,000 - \$49,999 | 21 | 20.8% |
| \$50,000 + | 24 | 23.8% |
| <u>Developed Environment (N=102)</u> |  |  |
| Suburban | 49 | 48.0% |
| Urban | 47 | 46.1% |
| Rural | 6 | 5.9% |
| <u>Marital Status (N=102)</u> |  |  |
| Single partner, married | 47 | 46.1% |
| Never married or partnered | 25 | 24.5% |
| Single partner, never married | 12 | 11.8% |
| Multiple committed partners | 11 | 10.8% |
| Separated, divorced, or widowed | 7 | 6.9% |
| <u>History of sexual behaviors</u> |  |  |
| Ever engaged in deep kissing (N=102) | 98 | 96.1% |
| Ever engaged in vaginal/front hole sex (N=97) | 95 | 97.9% |
| Ever engaged in oral sex (N=96) | 95 | 99.0% |
| Ever engaged in insertive anal sex (N=95) | 57 | 60.0% |
| Ever engaged in rimming/anilingus (N=100) | 43 | 43.0% |
| Ever engaged in mutual masturbation (N=100) | 89 | 89.0% |
| <u>Gender-affirming transition</u> |  |  |
| Have transitioned or are transitioning (N=102) | 77 | 75.5% |
| Ever taken hormones for transition (N=76) | 58 | 76.3% |
| Ever had surgery for transitions (N=77) | 32 | 41.6% |
| <u>Substance use (N=100)</u> |  |  |
| Current alcohol use | 75 | 75.0% |
| Ever cannabis use | 54 | 54.0% |
| Current cannabis use | 43 | 43.0% |
| Ever cigarette use | 24 | 24.0% |
| Current cigarette use | 8 | 8.0% |
| Ever used illegal drugs | 21 | 21.0% |
| Ever illicitly used prescription painkillers | 25 | 25.0% |
| <u>Medical history</u> |  |  |
| Ever pregnant (N=102) | 14 | 13.7% |
| Ever had a cervical Pap test (N=102) | 83 | 81.4% |
| Ever had an anal Pap test (N=102) | 6 | 5.9% |
| Ever had HPV (N=102) | 8 | 7.8% |
| Ever had other STI (N=100) | 22 | 22.0% |
| At least one dose of HPV vaccine (N=102) | 58 | 56.9% |
| <u>Medical Insurance</u> |  |  |
|  | 99 | - |
| Private insurance | 44 | 44.4% |
| Public insurance | 32 | 32.3% |
| Parents' private insurance | 23 | 23.2% |
| <u>Ever Delayed Checkups</u> |  |  |
|  | 102 | - |
| Yes, to avoid dysphoria or physical discomfort | 61 | 59.8% |
| Yes, fear of discrimination | 16 | 15.7% |
| Yes, provider lacked knowledge of trans-specific care | 6 | 5.9% |
| No | 19 | 18.6% |
| <u>Ever Delayed Treatment for Injury or Illness</u> | 102 | - |
| Yes, to avoid dysphoria or physical discomfort | 37 | 36.3% |
| Yes, fear of discrimination | 14 | 13.7% |
| Yes, provider lacked knowledge of trans-specific care | 3 | 2.9% |
| No | 48 | 47.1% |

Approximately half (48%) of participants identified as nonbinary, genderfluid, or agender and half (49%) as transgender male, transmasculine, or male (note that people who identify as male and were AFAB may or may not include being transgender as part of their gender identity). Most participants identified their sexual orientation as bisexual, pansexual, and omnisexual (43%) or simply as queer (36%). The vast majorities of participants reported a history of deep kissing (96%), mutual masturbation (89%), vaginal/front hole sex (98%), and oral sex (99%), while 60% reported ever engaging in anal sex, and 43% having ever engaged in anilingus/rimming. When asked about partners within the last 6 months, participants overall reported similar numbers of partners with a vagina and partners with a penis for vaginal, anal, or oral sex. A table of recent (past 6 month) sexual activity by sex type and partner genitals is given in the Supporting Information (Table S1).

About 75% of participants responded that they have transitioned or are currently transitioning from living as the gender assigned at birth, 76% of whom reported ever taking hormones as a part of gender-affirming medical care. Only 42% of transitioned or transitioning participants reported gender-affirming surgery. The vast majority (86%) of participants reported never being pregnant. Most participants indicated currently using alcohol (75%) and marijuana (54%), while only 32% reported ever smoking cigarettes. More than half (57%) reported receiving at least one dose of a vaccination against HPV, and most (81%) reported having ever had a cervical pap smear. While none of the participants reported being uninsured, 81% reported delaying regular medical checkups. Most of these (59%) were due to fear of physical discomfort or dysphoria. Most participants (53%) also reported delaying care for acute medical needs, with 36% of participants reporting delaying acute treatment due to concerns for physical discomfort or dysphoria.

### HPV prevalence and determinants

HPV prevalence varied across oral, cervicovaginal, and anal sites and across demographic and behavioral characteristics (Table 2). HPV was detected in 8.8% of valid tests on oral samples, 30.5% of valid tests on cervicovaginal samples, and 39.6% of valid tests on anal samples. High-risk HPV, which includes genotypes 16, 18, 31, 33, 35, 39, 45, 51, 52, 56, 58, and 59, was detected in 6.6% of oral, 26.8% of cervicovaginal, and 27.1% of anal samples. The genotype-specific HPV prevalence by site is given in the Supporting Information (Table S2).

**Table 2:**
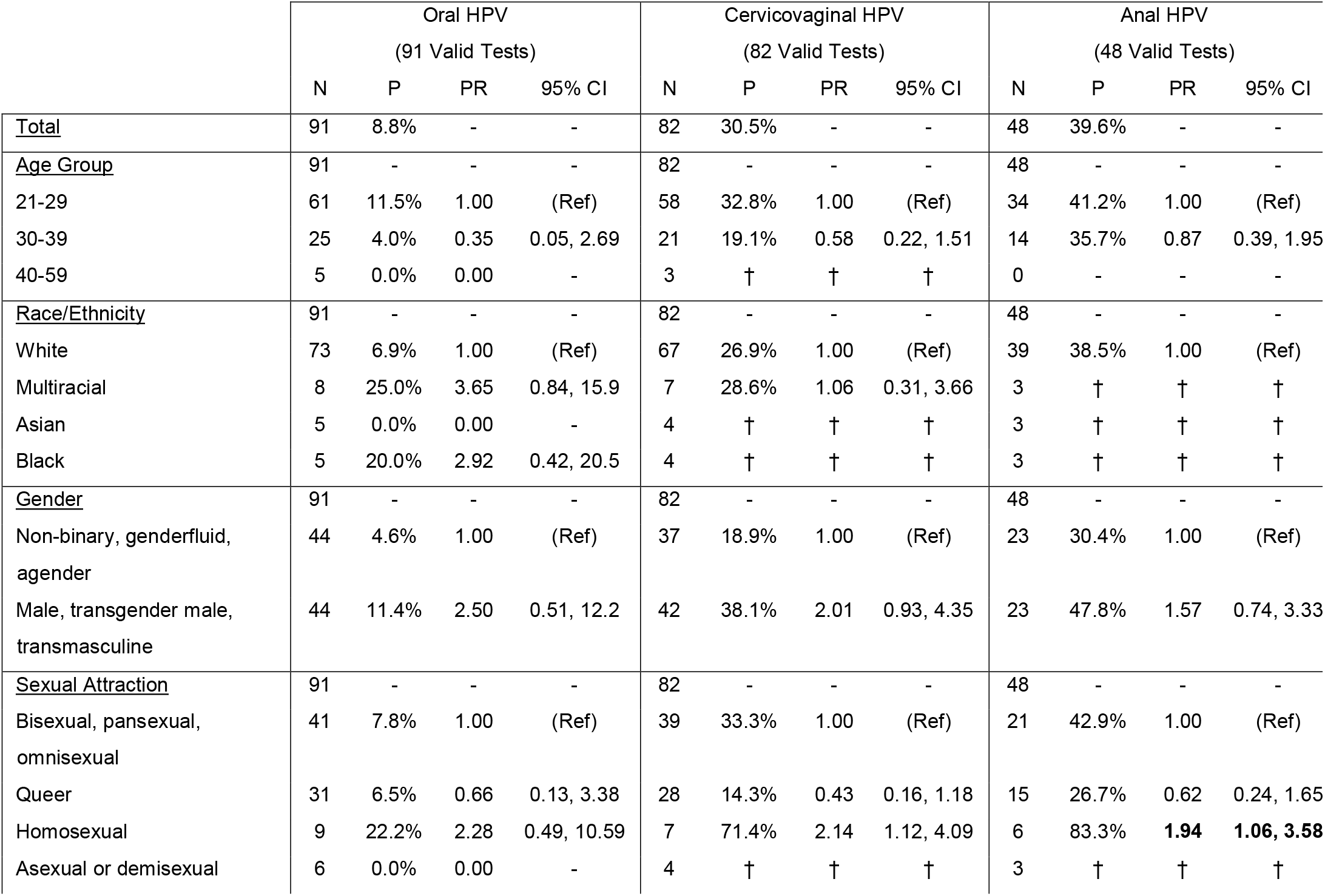
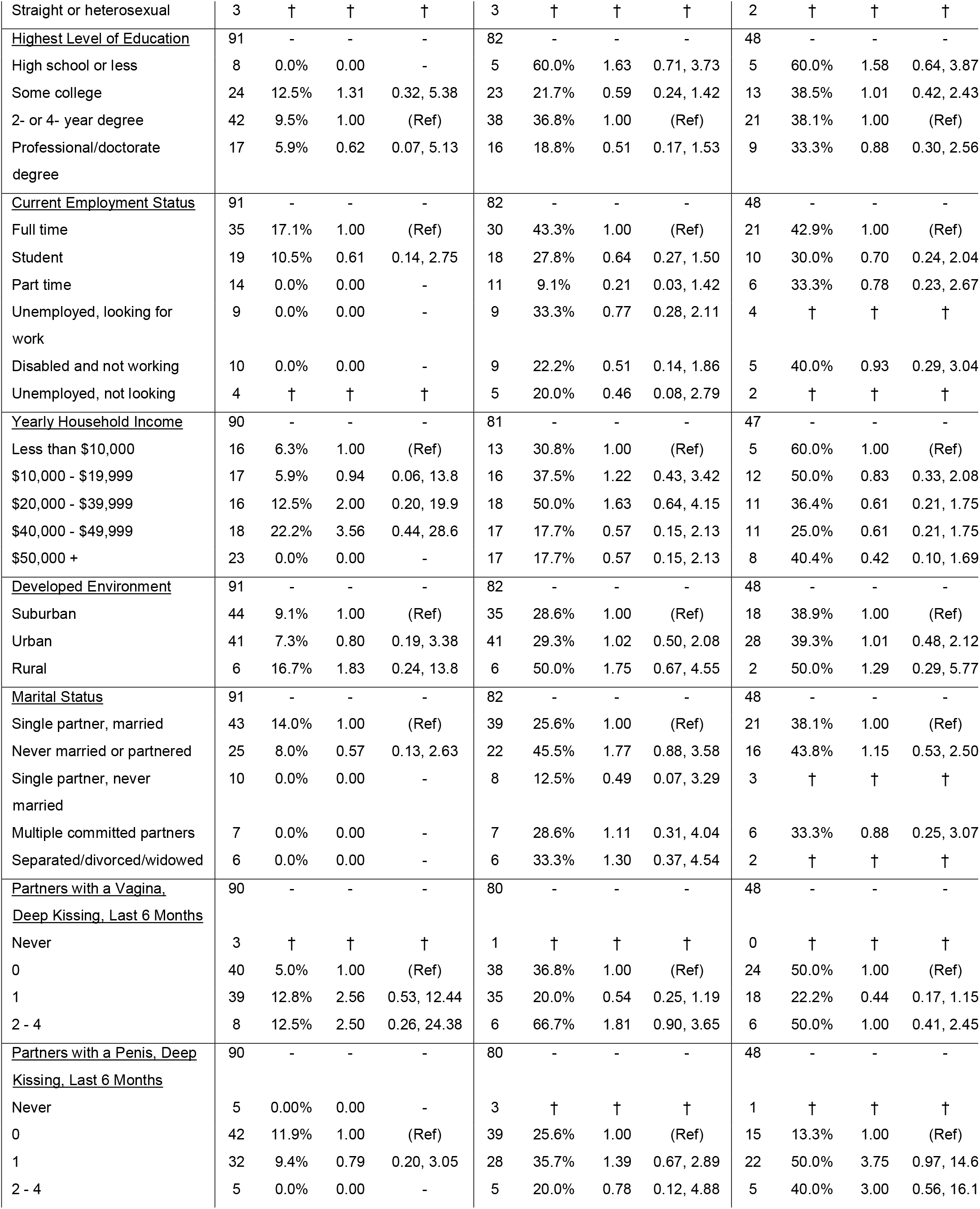
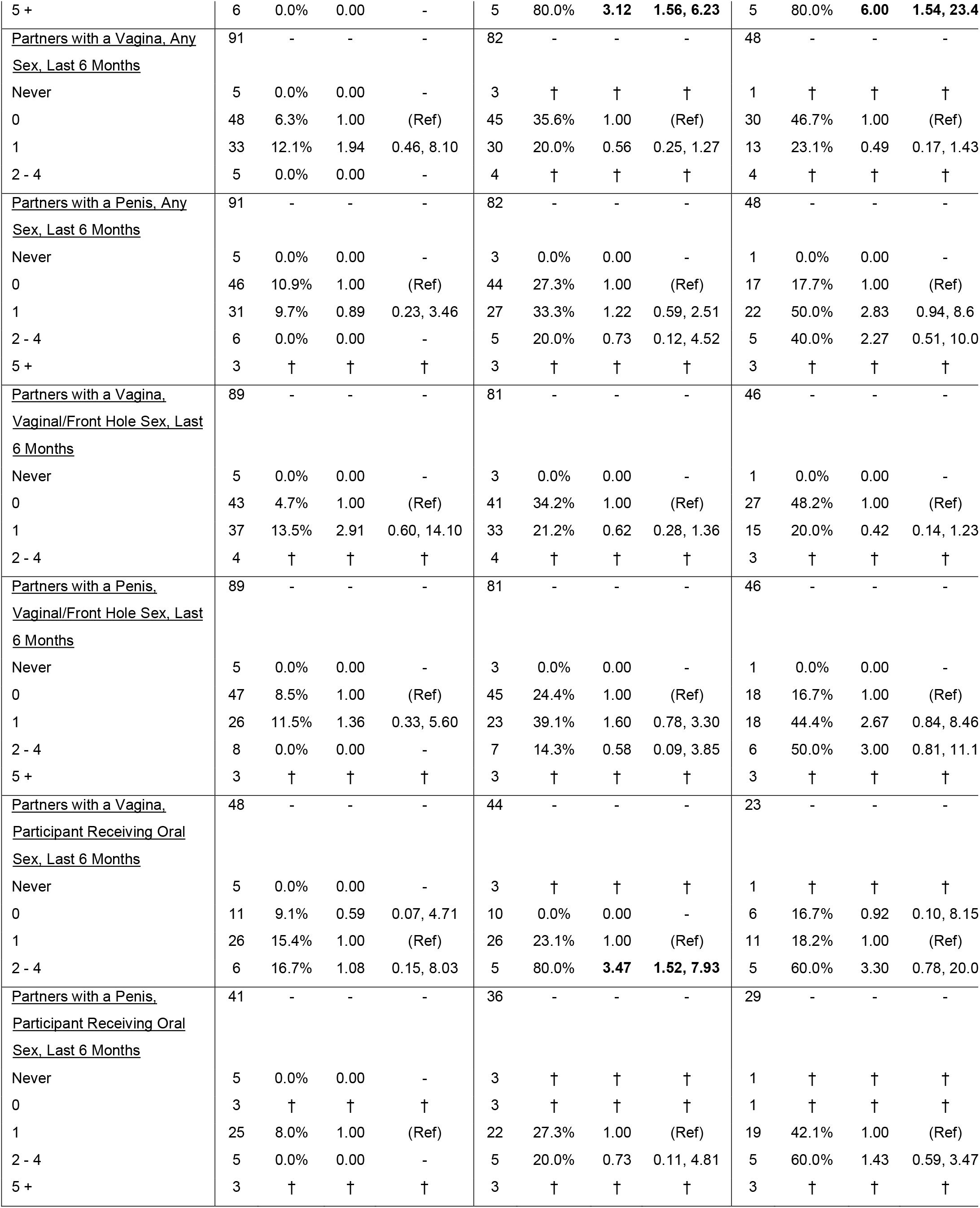
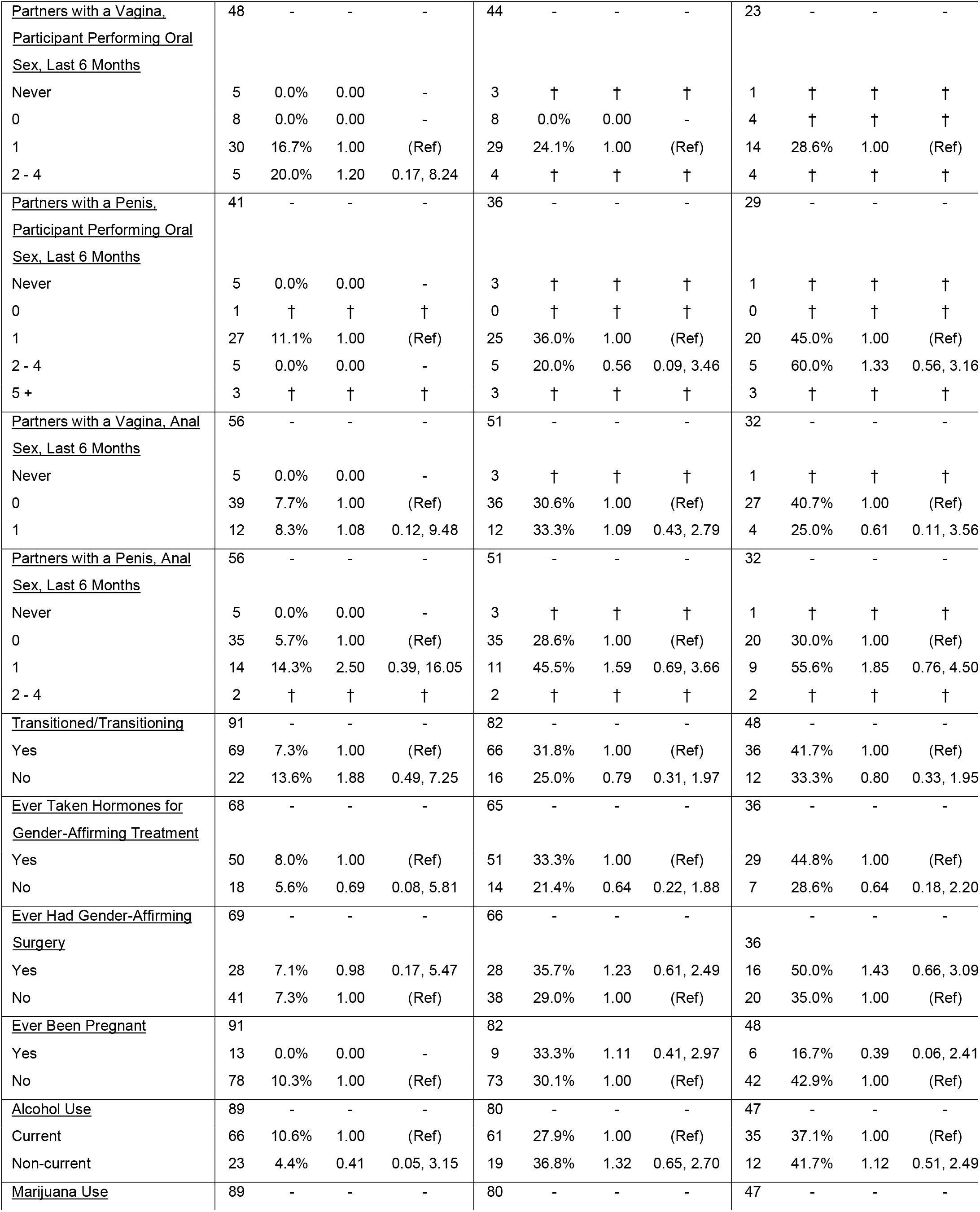
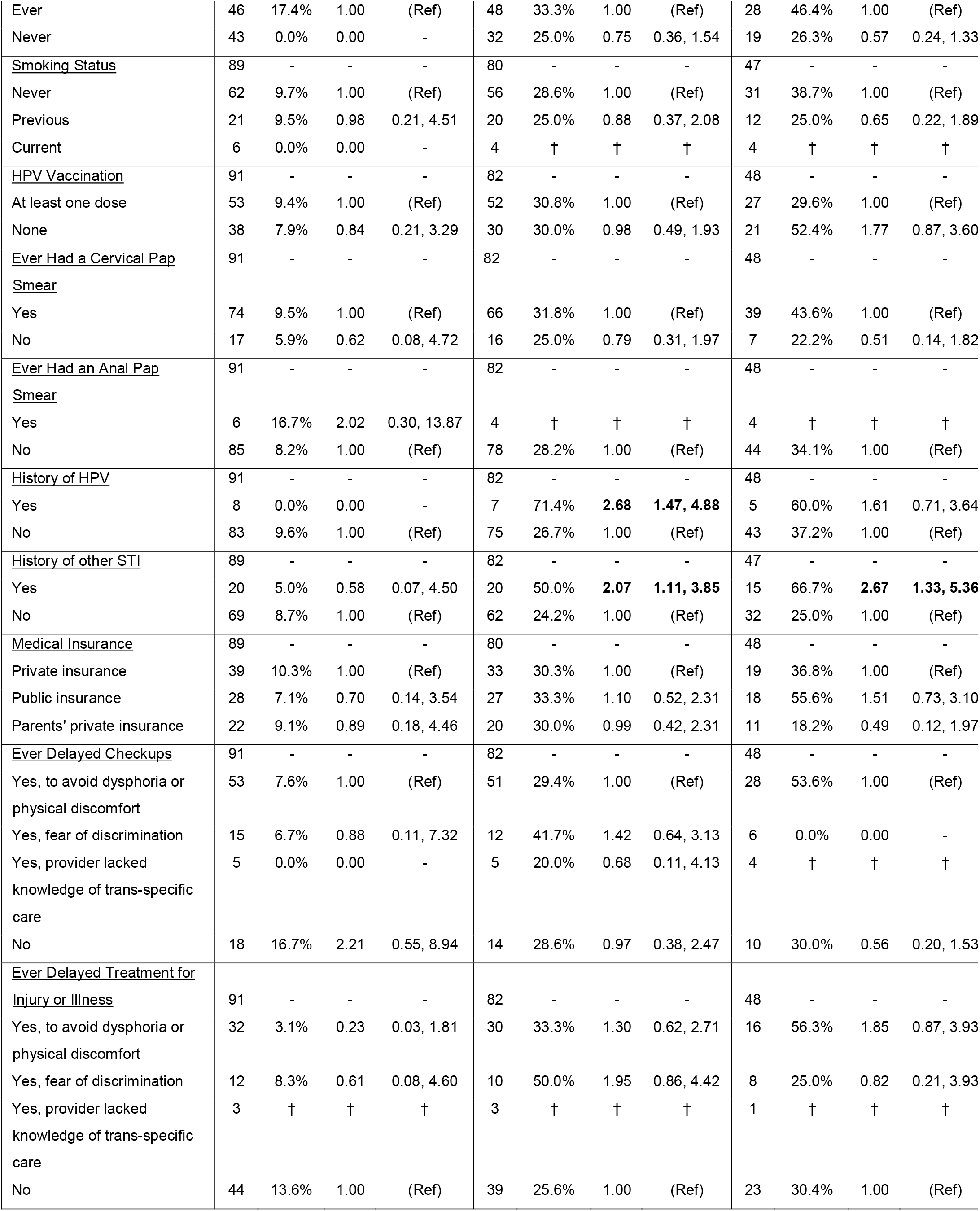
HPV prevalence and prevalence ratios by site and participant demographic, behavioral, and medical characteristics. Significantly different prevalence ratios are highlighted and bolded. N = number of individuals with valid test results in each group, P = prevalence of HPV within each group, PR = prevalence ratio of HPV in each group compared to referent group, and 95% CI = 95% confidence interval for each prevalence ratio. Bolded values are statistically significant at level of significance α=0.05. †Cells with fewer than 5 participants are censored.

Of the 44 participants with both valid cervicovaginal and anal test results, 25.0% of pairs of anal and cervicovaginal test results detected the presence of HPV of the same genotype. These concordant infections represented 34.4% of cervicovaginal infections in this group and 50.0% of anal infections. Of the 75 participants with valid results for both oral and cervicovaginal tests, 6.7% of pairs of oral and cervicovaginal tests detected the presence of the same genotype of HPV. These concordant infections represented only 12.2% of cervicovaginal infections in this group but 71.4% of oral infections. The HPV genotype-specific concordance for anal/cervicovaginal and oral/ cervicovaginal samples in the supporting information (Table S3).

No statistically significant associations were found between any characteristic and oral HPV infection. Cervicovaginal HPV infection was statistically significantly associated with the number of deep kissing partners with a penis in the past 6 months (PR 3.12 (95% CI: 1.56, 6.23) for 5+ partners vs 0 partners), number of partners with a vagina from whom the participant received oral sex (PR 3.47 (95% CI: 1.52, 7.93) for 2-4 partners vs 0 partners), a history of HPV (PR 2.68 (95%CI 1.47, 4.88)), and a history of other STI (PR 2.07 (95%CI 1.11, 3.85)). Anal HPV infection was statistically significantly associated with sexual attraction identity (PR 1.94 (95% CI: 1.06, 3.58) for identifying as homosexual vs bisexual, pansexual, or omnisexual), number of deep kissing partners with a penis in the past 6 months (PR 6.00 (95% CI: 1.54, 2.34) for 5+ partners vs 0 partners), and a history of other STI (PR 2.67 (95%CI 1.33, 5.36)).

## Discussion

We evaluated the prevalence of cervicovaginal, oral, and anal HPV in a Midwestern TGD AFAB population and investigated the potential association of infection with demographics, health factors, and sexual behaviors. We found HPV prevalence of 8.8% in oral samples, 30.5% in cervicovaginal samples, and 39.6% in anal samples. In a partner study using similar protocols for oral and cervicovaginal HPV detection in a general population with a similar age profile, 10.5% of cisgender women (and 9.0% of cisgender men) in the sample were positive for oral HPV, and 20% of cisgender women were positive for cervicovaginal HPV [32] (anal swabs were not collected in the partner study). This comparison suggests that the TGD AFAB population may have higher cervicovaginal HPV prevalence than but similar oral HPV prevalence to the cisgender women. A separate study in a TGD AFAB population, found 16% high-risk HPV prevalence in physician-collected cervicovaginal swabs, compared to the 26.8% high-risk HPV prevalence in our study [13]. Overall, we found no evidence that TGD people AFAB are at a lower risk for HPV and HPV-related cancers, and it is therefore important that screening for cervicovaginal HPV is accessible through affirming medical providers, or, potentially, at-home self-screening.

We found a high prevalence of anal HPV among valid tests (39.6%). A meta-analysis of anal HPV prevalence in cisgender women found that cisgender women without HPV-associated cervical pathology had anal HPV prevalence ranging from 4–22%, and prevalence in those with HPV-related cervical pathology ranged from 23–36% [33]. It is concerning, however, that despite modeling our anal sample collection instructions off an established protocol [29], we had low sample validity (that is, low detection of human control indicators) in our anal samples (47% valid). Validity was much higher among oral (89% valid) and cervicovaginal (80% valid) samples. Future work could involve instructional videos demonstrating anal (and cervicovaginal) sample collection techniques, and future work should consider whether there are particular challenges for anal sampling for TGD individuals. A follow-up study will also examine participants’ experiences with the cervicovaginal and anal sample self-collection.

We found significant differences in HPV prevalence across several participant characteristics and reported behavior. History of an STI diagnosis, likely a proxy for other behavior, was also associated with HPV at both cervicovaginal (PR: 2.07, 95%CI: 1.11, 3.85) and anal (PR: 2.67, 95%CI: 1.33, 5.36) sites. HPV is thought to be largely transmitted through sexual activity, and many previous studies have demonstrated associations in general populations [34–41]. Few associations rose to the level of statistical significance, with the exception of the number of deep kissing partners with a penis for both cervicovaginal and anal HPV infection and the number of partners with a vagina that a participant received oral sex from for cervicovaginal HPV infection. More broadly, in many sexual behavior categories, prevalence generally increased with the number of recent sexual partners, even if not statistically significantly. Additionally, although the effects were not statistically significant, there were indications that HPV prevalence among the participants identifying as male, transgender male, or transmasculine was higher than those identifying as nonbinary, genderfluid or agender. There were no significant differences in HPV prevalence by transition status, use of gender affirming hormones, or gender-affirming surgery.

We found a high degree of cervicovaginal–anal concordance among the participants with valid results for both anal and cervicovaginal tests. The high concordance of cervicovaginal–anal infection highlights the need to treat HPV as a multisite infection. One limitation is that we cannot tell from our data whether this correlation is the result of autoinoculation from site to site or separate transmission events from the same partner to different sites. In future work, the autoinoculation vs transmission explanations could be further evaluated using gene sequencing and mapping of samples from participants, or by utilizing contact tracing and partner testing. We found many oral infections were genotype concordant with cervicovaginal infections (71%), but few cervicovaginal infections were concordant with an oral infection (12%). These results are consistent with patterns of oral-cervicovaginal concordance in the National Health and Nutrition Examination Survey (NHANES) [42]. Together, our results are consistent with cervicovaginal infections being the source of anal and oral infections.

One strength of our study is the multisite design, incorporating oral, cervicovaginal, and anal self-sampling and HPV testing. Additionally, because many gender minority studies in the US are focused on urban, coastal populations, this Midwestern cohort helps to bring context and enhance the generalizability of gender minority studies. We also explicitly included nonbinary and gender diverse participants in this study, who are often overlooked in gender minority research. One limitation of our study is the small sample size, limiting statistical power, although other comparable studies have also had limited sample sizes [13]. Additionally, lack of racial and ethnic diversity in this pilot study is a strong limitation, as there may be important intersectional effects for TGD people of color. Indeed, we found high HPV prevalence among the non-White participants in this study. Future work should use targeted recruitment strategies to achieve better racial and ethnic representation. Finally, while the age distribution of the cohort (66% aged <30 years) is not inherently a limitation, it should be accounted for when try to generalize the results, since HPV prevalence varies by age [42].

## Conclusion

In our multisite (cervicovaginal, oral, anal) HPV sampling study in TGD people AFAB, we found HPV oral prevalence consistent with cisgender populations but higher cervicovaginal and anal infection. Our work suggests that the risk of HPV-related cancers in TGD people AFAB may be just as high, if not higher, than in cisgender women. It is essential that we reduce barriers to cancer screening for TGD populations, such as through the development of a clinically approved self-screening HPV test.

## Data Availability

All data produced in the present study are available upon reasonable request to the authors. A data use agreement will be required.

## Acknowledgments

We acknowledge our community consultants and our participants, without whom this work would not have been possible.

## Supporting Information

**Table S1:** Demographic, behavioral, and medical history characteristics of participants. N is the number of participants answering the question, and n is the number of respondents answering in the affirmative.

|  | n | % |
| --- | --- | --- |
| <u>Partners with a Vagina, Deep Kissing, Last 6 Months</u> | 99 | - |
| Never | 3 | 3.0% |
| 0 | 44 | 44.4% |
| 1 | 43 | 43.4% |
| 2 - 4 | 9 | 9.1% |
| 5 + | 0 | 0.0% |
| <u>Partners with a Penis, Deep Kissing, Last 6 Months</u> | 99 | - |
| Never | 5 | 5.1% |
| 0 | 49 | 49.5% |
| 1 | 33 | 33.3% |
| 2 - 4 | 6 | 6.1% |
| 5 + | 6 | 6.1% |
| <u>Partners with a Vagina, Vaginal / Oral / Anal Sex, Last 6 Months</u> | 102 | - |
| Never | 5 | 4.9% |
| 0 | 53 | 52.0% |
| 1 | 38 | 37.3% |
| 2 - 4 | 6 | 5.9% |
| 5 + | 0 | 0.0% |
| <u>Partners with a Penis, Vaginal / Oral / Anal Sex, Last 6 Months</u> | 102 | - |
| Never | 5 | 4.9% |
| 0 | 54 | 52.9% |
| 1 | 33 | 32.4% |
| 2 - 4 | 7 | 6.9% |
| 5 + | 3 | 2.9% |
| <u>Partners with a Vagina, Vaginal / Front Hole Sex, Last 6 Months</u> | 100 | - |
| Never | 5 | 5.0% |
| 0 | 49 | 49.0% |
| 1 | 41 | 41.0% |
| 2 - 4 | 5 | 5.0% |
| 5 + | 0 | 0.0% |
| <u>Partners with a Penis, Vaginal / Front Hole Sex, Last 6 Months</u> | 100 | - |
| Never | 5 | 5.0% |
| 0 | 55 | 55.0% |
| 1 | 28 | 28.0% |
| 2 - 4 | 9 | 9.0% |
| 5 + | 3 | 3.0% |
| <u>Partners with a Vagina, Participant Receiving Oral Sex, Last 6</u> | 55 | - |
| <u>Months</u> |  |  |
| Never | 5 | 9.1% |
| 0 | 12 | 21.8% |
| 1 | 31 | 56.6% |
| 2 - 4 | 7 | 12.7% |
| 5 + | 0 | 0.0% |
| <u>Partners with a Penis, Participant Receiving Oral Sex, Last 6</u> | 44 | - |
| <u>Months</u> |  |  |
| Never | 5 | 11.4% |
| 0 | 3 | 6.8% |
| 1 | 28 | 63.6% |
| 2 - 4 | 5 | 11.4% |
| 5 + | 3 | 6.8% |
| <u>Partners with a Vagina, Participant Performing Oral Sex, Last 6</u> | 55 | - |
| <u>Months</u> |  |  |
| Never | 5 | 9.1% |
| 0 | 10 | 18.2% |
| 1 | 34 | 61.8% |
| 2 - 4 | 6 | 10.9% |
| 5 + | 0 | 0.0% |
| <u>Partners with a Penis, Participant Performing Oral Sex, Last 6</u> | 44 | - |
| <u>Months</u> |  |  |
| Never | 5 | 11.4% |
| 0 | 2 | 4.6% |
| 1 | 28 | 63.6% |
| 2 - 4 | 6 | 13.6% |
| 5 + | 3 | 6.8% |
| <u>Partners with a Vagina, Anal Sex, Last 6 Months</u> | 62 | - |
| Never | 5 | 8.1% |
| 0 | 43 | 69.4% |
| 1 | 14 | 22.6% |
| 2 - 4 | 0 | 0.0% |
| 5 + | 0 | 0.0% |
| <u>Partners with a Penis, Anal Sex, Last 6 Months</u> | 62 | - |
| Never | 5 | 8.1% |
| 0 | 40 | 64.5% |
| 1 | 14 | 22.6% |
| 2 - 4 | 3 | 4.8% |
| 5 + | 0 | 0.0% |

**Table S2.** Prevalence of HPV genotypes in each sample type, and number and percentage of anal/ cervicovaginal and oral/ cervicovaginal matches.

| HPV Genotype | Oral<br>(91 Valid Tests) |  | Cervicovaginal (82<br>Valid Tests) |  | Anal<br>(48 Valid Tests) |  |
| --- | --- | --- | --- | --- | --- | --- |
|  | Number<br>Positive | % | Number<br>Positive | % | Number<br>Positive | % |
| Any | 8 | 8.8% | 25 | 30.5% | 16 | 33.3% |
| HPV 6 | 0 | 0.0% | 2 | 2.4% | 1 | 2.1% |
| HPV 11 | 0 | 0.0% | 0 | 0.0% | 0 | 0.0% |
| HPV 16 | 1 | 1.1% | 3 | 3.7% | 3 | 6.3% |
| HPV 18 | 2 | 2.2% | 5 | 6.1% | 1 | 2.1% |
| HPV 31 | 0 | 0.0% | 0 | 0.0% | 0 | 0.0% |
| HPV 33 | 0 | 0.0% | 0 | 0.0% | 1 | 2.1% |
| HPV 35 | 0 | 0.0% | 0 | 0.0% | 1 | 2.1% |
| HPV 39 | 1 | 1.1% | 3 | 3.7% | 3 | 6.3% |
| HPV 45 | 0 | 0.0% | 1 | 1.2% | 0 | 0.0% |
| HPV 51 | 1 | 1.1% | 6 | 7.3% | 4 | 8.3% |
| HPV 52 | 0 | 0.0% | 4 | 4.9% | 1 | 2.1% |
| HPV 56 | 1 | 1.1% | 5 | 6.1% | 3 | 6.3% |
| HPV 58 | 0 | 0.0% | 1 | 1.2% | 0 | 0.0% |
| HPV 59 | 0 | 0.0% | 5 | 6.1% | 1 | 2.1% |
| HPV 66 | 2 | 2.2% | 3 | 3.7% | 3 | 6.3% |
| HPV 68 | 0 | 0.0% | 3 | 3.7% | 0 | 0.0% |
| HPV 73 | 0 | 0.0% | 2 | 2.4% | 0 | 0.0% |
| HPV 90 | 0 | 0.0% | 1 | 1.2% | 1 | 2.1% |
| COBAS Test (HPV 16, 18, 31, 33, 35, 39, 45, 51, 52, 56, 58, 59, 66, and 68) | 8 | 8.8% | 23 | 28.1% | 15 | 31.3% |
| IARC Group 1 (Carcinogenic) (HPV 16, 18, 31, 33, 35, 39, 45, 51, 52, 56, 58, and 59) | 6 | 6.6% | 22 | 26.8% | 13 | 27.1% |
| Included in Gardasil (HPV 6, 11, 16, and 18) | 3 | 3.3% | 10 | 12.2% | 5 | 10.4% |
| Included in Gardasil 9 (HPV 6, 11, 16, 18, 31, 33, 45, 52, and 58) | 3 | 3.3% | 14 | 17.1% | 6 | 12.5% |
| Cause Genital Warts (HPV 6 and 11) | 0 | 0.0% | 2 | 2.4% | 1 | 2.1% |

**Table S3.** Genotype concordance between anal and cervicovaginal and between oral and cervicovaginal HPV infections.

| HPV Genotype | Anal/cervicovaginal genotype matches<br>(44 participants with valid tests for both samples) |  |  | Oral/cervicovaginal genotype matches<br>(75 participants with valid tests for both samples) |  |  |
| --- | --- | --- | --- | --- | --- | --- |
|  | Number positive<br>for both | % of positive<br>cervicovaginal tests | % of positive<br>anal Tests | Number positive<br>for both | % of positive<br>cervicovaginal tests | % of positive<br>oral tests |
| Any | 11 | 34.4% | 50.0% | 5 | 12.2% | 71.4% |
| HPV 6 | 1 | 50.0% | 100.0% | 0 | 0.0% | - |
| HPV 11 | 0 | - | - | 0 | - | - |
| HPV 16 | 1 | 33.3% | 33.3% | 0 | 0.0% | 0.0% |
| HPV 18 | 0 | 0.0% | 0.0% | 1 | 25.0% | 50.0% |
| HPV 31 | 0 | - | - | 0 | - | - |
| HPV 33 | 0 | - | 0.0% | 0 | - | - |
| HPV 35 | 0 | - | 0.0% | 0 | - | - |
| HPV 39 | 2 | 100.0% | 66.7% | 0 | 0.0% | - |
| HPV 45 | 0 | 0.0% | - | 0 | 0.0% | - |
| HPV 51 | 1 | 33.3% | 25.0% | 1 | 16.7% | 100.0% |
| HPV 52 | 1 | 25.0% | 100.0% | 0 | 0.0% | - |
| HPV 56 | 2 | 50.0% | 66.7% | 1 | 20.0% | 100.0% |
| HPV 58 | 0 | 0.0% | - | 0 | 0.0% | - |
| HPV 59 | 1 | 25.0% | 100.0% | 0 | 0.0% | - |
| HPV 66 | 1 | 100.0% | 50.0% | 2 | 66.7% | 100.0% |
| HPV 68 | 0 | 0.0% | - | 0 | 0.0% | - |
| HPV 73 | 0 | 0.0% | - | 0 | 0.0% | - |
| HPV 90 | 1 | 100.0% | 100.0% | 0 | 0.0% | - |

## References

[1] Center for Disease Control and Prevention. Fact Sheet: United States Cancer Statistics Data Brief, Cancers Associated with Human Papillomavirus, United States, 2011 to 2015. USCS Data Brief. 2018;4.

[2] Curry SJ, Krist AH, Owens DK, Barry MJ, Caughey AB, Davidson KW, et al. Screening for Cervical Cancer. JAMA. 2018;320(7):674.

[3] Reisner SL, Murchison GR. A global research synthesis of HIV and STI biobehavioural risks in female-to-male transgender adults. Global Public Health. 2016;11(7-8):866–87.

[4] Agenor M, Peitzmeier SM, Bernstein IM, McDowell M, Alizaga NM, Reisner SL, et al. Perceptions of cervical cancer risk and screening among transmasculine individuals: patient and provider perspectives. Culture, Health & Sexuality. 2016;18(10):1192–206.

[5] Peitzmeier SM, Reisner SL, Harigopal P, Potter J. Female-to-male patients have high prevalence of unsatisfactory paps compared to non-transgender females: Implications for cervical cancer screening. Journal of General Internal Medicine. 2014;29(5):778–84.

[6] Gatos KC. A Literature Review of Cervical Cancer Screening in Transgender Men. Nursing for Women’s Health. 2018;22(1):52–62.

[7] Obedin-Maliver J, de Haan G. Gynecologic Care for Transgender Adults. Current Obstetrics and Gynecology Reports. 2017;6(2):140–8.

[8] Seay J, Ranck A, Weiss R, Salgado C, Fein L, Kobetz E. Understanding Transgender Men’s Experiences with and Preferences for Cervical Cancer Screening: A Rapid Assessment Survey. LGBT Health. 2017;4(4):304–9.

[9] Potter J, Peitzmeier SM, Bernstein I, Reisner SL, Alizaga NM, Aǵenor M, et al. Cervical Cancer Screening for Patients on the Female-to-Male Spectrum: a Narrative Review and Guide for Clinicians. Journal of General Internal Medicine. 2015;30(12):1857–64.

[10] Peitzmeier SM, Khullar K, Reisner SL, Potter J. Pap test use is lower among female-to-male patients than nontransgender women. American Journal of Preventive Medicine. 2014;47(6):808–12.

[11] Dhillon N, Oliffe JL, Kelly MT, Krist J. Bridging barriers to cervical cancer screening in transgender men: a scoping review. American journal of men’s health. 2020;14(3):1557988320925691.

[12] Kattari SK, Gross EB, Harner V, Andrus E, Stroumsa D, Moravek MB, et al. “Doing It on My Own Terms”: Transgender and Nonbinary Adults’ Experiences with HPV Self-Swabbing Home Testing Kits. Women’s Reproductive Health. 2022:1–17.

[13] Reisner SL, Deutsch MB, Peitzmeier SM, White Hughto JM, Cavanaugh TP, Pardee DJ, et al. Test performance and acceptability of self-versus provider-collected swabs for high-risk HPV DNA testing in female-to-male trans masculine patients. PLoS ONE. 2018;13(3):1–21.

[14] Reisner SL, Deutsch MB, Peitzmeier SM, White Hughto JM, Cavanaugh T, Pardee DJ, et al. Comparing self- and provider-collected swabbing for HPV DNA testing in female-to-male transgender adult patients: A mixed-methods biobehavioral study protocol. BMC Infectious Diseases. 2017;17(1):1–10.

[15] McDowell M, Pardee DJ, Peitzmeier S, Reisner SL, Aǵenor M, Alizaga N, et al. Cervical Cancer Screening Preferences Among Trans-Masculine Individuals: Patient-Collected Human Papillomavirus Vaginal Swabs Versus Provider-Administered Pap Tests. LGBT Health. 2017;4(4):252–9.

[16] Maza M, Meĺendez M, Herrera A, Hernandez X, Rodŕ ŕguez B, Soler M, et al. Cervical cancer screening with human papillomavirus self-sampling among transgender men in El Salvador. LGBT health. 2020;7(4):174–81.

[17] Brown B, Poteat T, Marg L, Galea JT. Human Papillomavirus-Related Cancer Surveillance, Prevention, and Screening among Transgender Men and Women: Neglected Populations at High Risk. LGBT Health. 2017;4(5):315–9.

[18] Jemal A, Simard EP, Dorell C, Noone AM, Markowitz LE, Kohler B, et al. Annual Report to the Nation on the Status of Cancer, 1975-2009, Featuring the Burden and Trends in Human Papillomavirus (HPV)-Associated Cancers and HPV Vaccination Coverage Levels. Journal of the National Cancer Institute. 2013;105(3):175–201.

[19] Siegel RL, Miller KD, Jemal A. Cancer statistics, 2018. CA: A Cancer Journal for Clinicians. 2018;68(1):7–30.

[20] Van Dyne EA, Henley SJ, Saraiya M, Thomas CC, Markowitz LE, Benard VB. Trends in Human Papillomavirus–Associated Cancers — United States, 1999–2015. Morbidity and Mortality Weekly Report. 2018;67(33):918–24.

[21] Steinau M, Hariri S, Gillison ML, Broutian TR, Dunne EF, Tong ZyY, et al. Prevalence of Cervical and Oral Human Papillomavirus Infections Among US Women. The Journal of Infectious Diseases. 2014;209(11):1739–43.

[22] Brouwer AF, Eisenberg MC, Carey TE, Meza R. Trends in HPV cervical and seroprevalence and associations between oral and genital infection and serum antibodies in NHANES 2003-2012. BMC Infectious Diseases. 2015;15(1):575.

[23] Kedarisetty S, Orosco RK, Hecht AS, Chang DC, Weissbrod PA, Coffey CS. Concordant Oral and Vaginal Human Papillomavirus Infection in the United States. JAMA Otolaryngology–Head & Neck Surgery. 2016;8895:1–9.

[24] Brouwer AF, Meza R, Eisenberg MC. Transmission heterogeneity and autoinoculation in a multisite infection model of HPV. Mathematical Biosciences. 2015;270:115–25.

[25] Patel EU, Rositch AF, Gravitt PE, Tobian AAR. Concordance of Penile and Oral Human Papillomavirus Infections Among Men in the United States. The Journal of Infectious Diseases. 2017;215(8):1207–11.

[26] Sonawane K, Suk R, Chiao EY, Chhatwal J, Qiu P, Wilkin T, et al. Oral human papillomavirus infection: Differences in prevalence between sexes and concordance with genital human papillomavirus infection, NHANES 2011 to 2014. Annals of Internal Medicine. 2017;167(10):714–24.

[27] Rovers Medical Devices. Evalyn®Brush; 2022. https://www.roversmedicaldevices.com/cell-samplingdevices/evalyn-brush/. Accessed February 5, 2022.

[28] Ejegod DM, Pedersen H, Alzua GP, Pedersen C, Bonde J. Time and temperature dependent analytical stability of dry-collected Evalyn HPV self-sampling brush for cervical cancer screening. Papillomavirus Research. 2018;5:192200.

[29] Meites E, Gorbach PM, Gratzer B, Panicker G, Steinau M, Collins T, et al. Monitoring for human papillomavirus vaccine impact among gay, bisexual, and other men who have sex with men—United States, 2012–2014. The Journal of infectious diseases. 2016;214(5):689–96.

[30] Eisenberg MC, Campredon LP, Brouwer AF, Walline HM, Marinelli BM, Lau YK, et al. Dynamics and Determinants of HPV Infection: The Michigan HPV and Oropharyngeal Cancer (M-HOC) Study. BMJ Open. 2018;8(10):e021618.

[31] Walline HM, Komarck C, McHugh JB, Byrd SA, Spector ME, Hauff SJ, et al. High-risk human papillomavirus detection in oropharyngeal, nasopharyngeal, and oral cavity cancers: comparison of multiple methods. JAMA Otolaryngology–Head & Neck Surgery. 2013;139(12):1320–7.

[32] Brouwer AF, Campredon LP, Walline HM, Marinelli BM, Goudsmit CM, Thomas TB, et al. Prevalence and determinants of oral and cervicogenital HPV infection: Baseline analysis of the Michigan HPV and Oropharyngeal Cancer (MHOC) cohort study. Plos one. 2022;17(5):e0268104.

[33] Stier EA, Sebring MC, Mendez AE, Ba FS, Trimble DD, Chiao EY. Prevalence of anal human papillomavirus infection and anal HPV-related disorders in women: a systematic review. American journal of obstetrics and gynecology. 2015;213(3):278–309.

[34] Cook RL, Thompson EL, Kelso NE, Friary J, Hosford J, Barkley P, et al. Sexual behaviors and other risk factors for oral human papillomavirus infections in young women. Sexually Transmitted Diseases. 2014;41(8):486.

[35] Pickard RK, Xiao W, Broutian TR, He X, Gillison ML. The prevalence and incidence of oral human papillomavirus infection among young men and women, aged 18-30 years. Sexually Transmitted Diseases. 2012:559–66.

[36] Klein TW, Newton C, Larsen K, Lu L, Perkins I, Nong L, et al. The cannabinoid system and immune modulation. Journal of Leukocyte Biology. 2003;74(4):486–96.

[37] Brouwer AF, Campredon LP, Walline HM, Marinelli BM, Goudsmit CM, Thomas TB, et al. Incidence and clearance of oral and cervicogenital HPV infection: longitudinal analysis of the MHOC cohort study. BMJ Open. 2022;12(1):e056502.

[38] Edelstein ZR, Schwartz SM, Hawes S, Hughes JP, Feng Q, Stern ME, et al. Rates and determinants of oral human papillomavirus (HPV) infection in young men. Sexually Transmitted Diseases. 2012;39(11):860.

[39] D’Souza G, Cullen K, Bowie J, Thorpe R, Fakhry C. Differences in oral sexual behaviors by gender, age, and race explain observed differences in prevalence of oral human papillomavirus infection. PloS one. 2014;9(1):e86023.

[40] D’Souza G, Wentz A, Kluz N, Zhang Y, Sugar E, Youngfellow RM, et al. Sex differences in risk factors and natural history of oral human papillomavirus infection. The Journal of Infectious Diseases. 2016;213(12):1893–6.

[41] Chung CH, Bagheri A, D’Souza G. Epidemiology of oral human papillomavirus infection. Oral oncology. 2014;50(5):364–9.

[42] Brouwer AF, Eisenberg MC, Carey TE, Meza R. Multisite HPV infections in the United States (NHANES 2003– 2014): an overview and synthesis. Preventive medicine. 2019;123:288–98.

